# Age-stratified model of the COVID-19 epidemic to analyze the impact of relaxing lockdown measures: nowcasting and forecasting for Switzerland

**DOI:** 10.1101/2020.05.08.20095059

**Authors:** F. Balabdaoui, D. Mohr

**Affiliations:** Seminar of Statistics, Department of Mathematics, Swiss Federal Institute of Technology (ETH), Rämistrasse 101, Zurich, 8092, Switzerland; Department of Mechanical and Process Engineering, Swiss Federal Institute of Technology (ETH), Tannenstrasse 3, Zurich, 8092, Switzerland

**Keywords:** COVID-19, epidemic modeling, contact patterns, reproduction number, fatality ratio

## Abstract

Compartmental models enable the analysis and prediction of an epidemic including the number of infected, hospitalized and deceased individuals in a population. They allow for computational case studies on non-pharmaceutical interventions thereby providing an important basis for policy makers. While research is ongoing on the transmission dynamics of the SARS-CoV-2 coronavirus, it is important to come up with epidemic models that can describe the main stages of the progression of the associated COVID-19 respiratory disease. We propose an age-stratified discrete compartment model as an alternative to differential equation based S-I-R type of models. The model captures the highly age-dependent progression of COVID-19 and is able to describe the day-by-day advancement of an infected individual in a modern health care system. The fully-identified model for Switzerland not only predicts the overall histories of the number of infected, hospitalized and deceased, but also the corresponding age-distributions. The model-based analysis of the outbreak reveals an average infection fatality ratio of 0.4% with a pronounced maximum of 9.5% for those aged ≥80 years. The predictions for different scenarios of relaxing the soft lockdown indicate a low risk of overloading the hospitals through a second wave of infections. However, there is a hidden risk of a significant increase in the total fatalities (by up to 200%) in case schools reopen with insufficient containment measures in place.

---

The coronavirus-induced COVID-19 epidemic^1^ is constantly pushing governments to take rapid decisions on measures for protecting the public health while minimizing economic damage. The emergency to act after exceeding infection rates of 1 out of 10’000 individuals has been recognized by most governments. The coronavirus containment measures often started by recommending social distancing and improved hand hygiene and ended by the complete lockdown of countries in extreme cases. After putting a country into an extraordinary state in response to a first epidemic wave, the next challenge for governments is the timely release of drastic measures to reduce the psychological and economic damage while preventing a possible second epidemic wave. Timely decision making is crucial when implementing and relaxing measures. During those two phases, there is a competition between avoiding COVID-19 related fatalities and preventing harm (and secondary casualties) due to economic recession.

Epidemic models^2^ provide an important mathematical tool to support decision making. A prominent example are the simulations performed by Ferguson et al.^3^ which (among others^4-6^) provided convincing evidence in favor of implementing strong non-pharmaceutical interventions in response to the COVID-19 outbreak. The S-I-R compartmental models^7^ divide a population into groups of susceptible (S), infected (I) and recovered (R) individuals. Adding more compartments allows for a refined description of specific epidemics. Such models range from SEIR^6,8,9^ and SUQC^10^ models of COVID-19 to models as complex as the SIDARTHE^11^ model which considers susceptible, infected, diagnosed, ailing, recognized, threatened, healed and extinct compartments. An important feature of COVID-19 is its highly non-uniform attack of the different age strata of society. Statistical analysis of data collected during the COVID-19 epidemic in Hubei^12^ reveals that the infection fatality ratio for individuals older than 80 is likely to be one order of magnitude higher than that for individuals of 50 years and younger. Age-stratified epidemic models are therefore particularly relevant when estimating the hospital load and fatalities related to COVID-19. Moreover, the age-dependent patterns of social contacts may be incorporated into age-stratified models. As a result, the obtained mathematical models provide not only estimates of the overall dynamics of an epidemic, but they are also able to predict the effect of age-dependent relaxation measures such as reopening school.

Here, we propose a novel approach to epidemic modeling to capture the age-dependent dynamics of COVID-19. Instead of using SIR-type of differential equations to describe the transfer between neighboring compartments, a discrete compartment model (Fig. 1) is built which mimics the different “trajectories” of individuals from exposure to healing or death. Aside from the standard compartments for susceptible and exposed individuals, the model differentiates between symptomatic and asymptomatic infected individuals.

**Fig 1.**
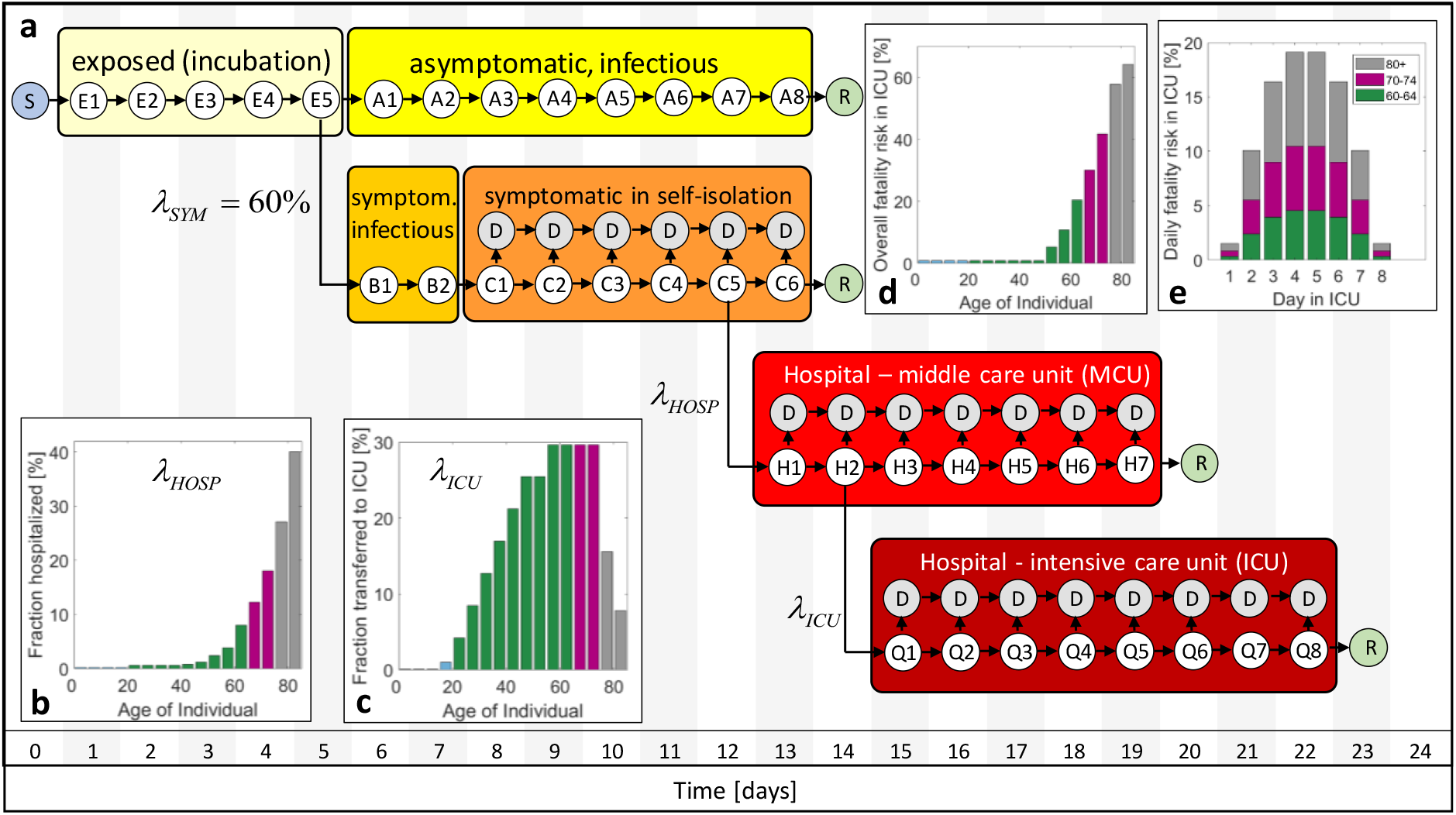
Discrete transmission model for COVID-19. **(a)** The different compartments comprise susceptible (S), exposed (E), asymptomatic (A), symptomatic before (B) and in self-isolation (C), hospitalized in MCU (H) and ICU (Q), removed (R) and deceased (D) individuals. Individuals are classified into sub-compartments E1, E2, etc. according to the number of days they have spent in a given comportment. **(b)** age-dependent probability of hospitalization of symptomatic individuals, **(c)** probability of transfer from MCU to ICU, **(d)** fatality risk in ICU, **(e)** daily fatality ratio in ICU exemplarily shown for age-groups 80+, 70-74 and 60-64.

The group of symptomatic is further split into a compartment of self-isolated individuals and those requiring hospitalization and admission to middle care (MCU) and intensive care units (ICU). Defining the first day of infection as Day 1, distinct sub-compartments are defined for all subsequent days until healing or death. The model is then updated on a daily basis by moving individuals to a specific sub-compartment for the subsequent day. Those moves are defined through shifting laws which account for the age-dependent probabilities of infection, admission to hospital, transfer to intensive care and death. As detailed in the Methods Section, the sizes of the compartments are set by the incubation time (5 days), the duration from the onset of symptoms to self-isolation (2 days), the average duration of viral shedding by asymptomatic individuals (8 days), the average duration spent in hospital (7 days) and in intensive care (8 days). In Fig. 1a, each encircled variable (E1, E2, etc) represents a vector whose components corresponds to the number of individuals in a certain age-group that are currently in a given sub-compartment (e.g. H2 for second day in hospital). In total, we differentiate among 16 age-groups from 0 to 79 years, plus age-group #17 which represents all individuals aged 80 years and over.

In addition to setting the time scales, the transfer probabilities need to be selected. Based on the results from the testing of small populations without any symptom-based pre-selection^13, 14, 15^, we assume that 60% of the infected develop symptoms irrespective of age. All other probability functions determining the model are expected to be partly country-specific. For example, the probability of infection depends on the social contact patterns. Also the probability of admission to hospital and ICU depends on the healthcare system and the local culture (e.g. specific guidelines for physicians, elderly refusing treatments that prolong life, availability of medical care options at retiring homes). The probability that an individual of a first age-group infects a susceptible individual of a second age-group is related to the number of contacts per day at home, at work, at school and at other locations (see contact maps depicted in Supplementary Fig. 1). Furthermore, it depends on the probability of transmission per contact which is inferred from the reproduction number of the epidemic before applying emergency measures.

Due to its high rate of infection per capita and the expected good mixing of infected and susceptible individuals in a small country, we chose Switzerland as a first example for illustrating the merits of the proposed epidemic model. Even though Switzerland ranks among the countries with the highest COVID-19 PCR testing rates per capita, the symptomatic patient bias in the reported number of cases is still too strong for model identification. We thus make use of the reported number of currently hospitalized and the accumulated number of deaths along with selected statistics on the age distributions. The age-dependency of the MCU to ICU transfer probability (Fig. 1c) is inferred from the age distributions in hospital and ICU in the canton Vaud (Supplementary Fig. 3). While up to 30% of the patients admitted to hospital will be transferred for age-groups 55 to 74 years, a significantly lower fraction of the individuals older than 70 years is transferred. The fatality risk in ICU (Fig. 1d) is estimated from the reported data after assuming that all deaths of individuals aged <75 years occur in ICU. Here, we observe a fatality risk of 40% in the age-group 70-74 years which appears to be low when compared to data reported for UK hospitals^16^. On the other hand, the canton Vaud reports that as many as 65% of all COVID-19 deaths occurs outside the hospital. We thus also allow for deaths of symptomatic patients in self-isolation with a probability of 2.3% and 7.3% for the age-groups 75-79 and ≥80 years, respectively. For the latter age-group, death is also assumed to occur in MCU with a probability of 18%. After setting the overall probability of deaths per compartment and age-group, a bump function is employed (Supplementary Information) to determine the daily fatality risks (Fig. 1e). Using the identified functions, the overall history of fatalities (Fig. 2a) and the age distribution of the deceased (Fig. 2b) are predicted with reasonable accuracy. It is worth noting that about 70% of all COVID-19 related deaths in Switzerland are individuals aged 80 years or older, while the fraction of deceased is below 0.1% for those aged <50 years. The age-dependency of the probability of hospitalization for symptomatic individuals (Fig. 1b) is estimated based on the reported age-distribution in hospital. The resulting function *λ_HOSP_* is then adjusted such that the model predicts the documented history (Fig. 2c) and age-distributions of the individuals in hospital (Fig. 2d) with good accuracy. The resulting probabilities of hospitalization per age-group fall into the 95% credible intervals for COVID-19 hospitalization estimated based on data for mainland China^17^.

**Fig 2.**
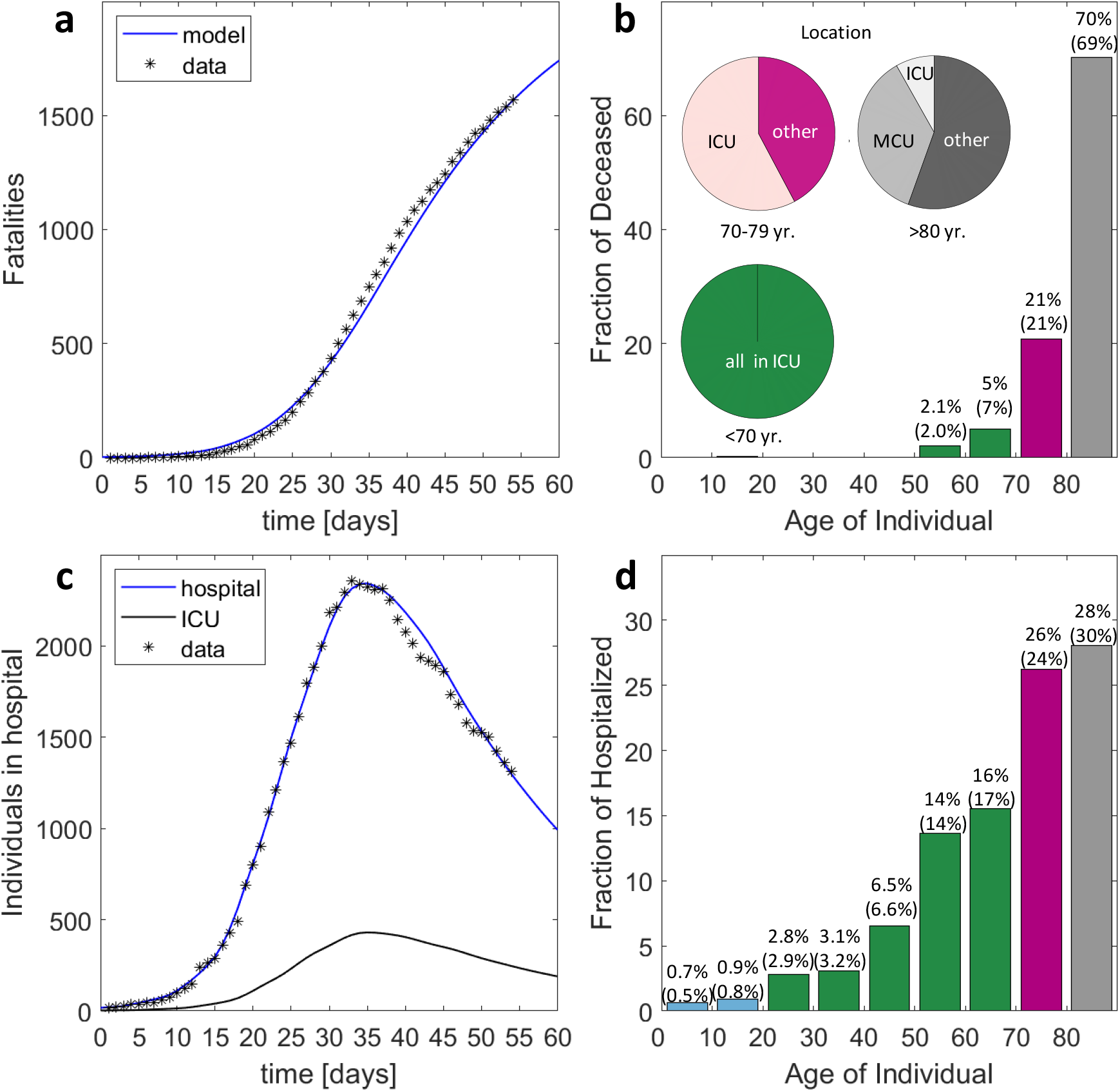
Model identification and validation for Switzerland. **(a)** History of fatalities, **(b)** age distribution of deceased and location distribution (pie charts), **(c)** history of individuals in hospital (MCU and ICU combined) and those in ICU, (d) age distribution of hospitalized. The values provided in parentheses in (b) and (d) correspond to the documented data for April 20. Day 1 on the charts shown in (a) and (c) corresponds to March 1, 2020.

A logarithmic plot of the history of hospitalizations shows a continuous change in slope after March 20^th^ (Supplementary Fig. 4) which is tentatively attributed to the progressive implementation of social distancing and improved hygiene. Due to the time lag of 13 days between first exposure and possible hospitalization (Fig. 1a), the rate of transmission is continuously reduced from March 7 onwards. On March 13, the government then advised to stay (and work) at home, closed schools and banned most private and public events. This (soft) lockdown not only changed the probabilities of transmission per contact (coefficients *β^*^* in our model, see Methods Section), but also the contact patterns between susceptible and infectious individuals (weights (*ϕ*. in our model). The ultimate result is an extraordinary state which leads to a decrease in the number of individuals in hospital after April 5. According to the model, the reproduction number (Fig. 3a) first decreases from 5 to 2.4 due to social distancing, before further decreasing from 2.4 to 0.7 due to the stay home policy and reduction of contacts at other locations. It is worth noting that the predicted average reproduction number for the extraordinary state falls into the confidence interval for estimates based on reported data^18^. The contact patterns before the extraordinary state result in a highly non-uniform spread of COVID-19. Among the exposed, the groups 10-14 and 15-19 are the ones showing the highest infection ratios, while older age-groups appear to be protected by the contact patterns among age-groups in Switzerland. The age distribution of the exposed becomes more uniform after March 13 (Figs. 3b-d) mostly due to school closure and reduced presence at work. The age-stratified model predicts significant differences in the reproduction numbers among age-groups (Figs. 3e-g) with a max-to-min ratio of 3.6 to 0.6 on March 13 and of 0.9 to 0.3 two weeks later. It is worth noting that the reproduction number for those aged 65 years and over is always lower than 1. The non-uniformity of the attack of COVID-19 is even more pronounced when evaluating the infection fatality ratio (IFR). The trained model predicts an IFR of 9.5% for the individuals older than 80 years (Fig. 4). It drops to 2.8% and 1.3% for the next two lower age-groups (75-79 and 70-74 years). Among the individuals in their sixties, there is still the risk of one death among 100 infected. An average IFR of about 0.38% is obtained when averaging over the whole age distribution of the infected. The estimated IFR for the 80+ age-group falls into the confidence interval for the IFR estimated based on data for mainland China^12^, while the IFRs for Switzerland are lower for all other age-groups.

**Fig 3.**
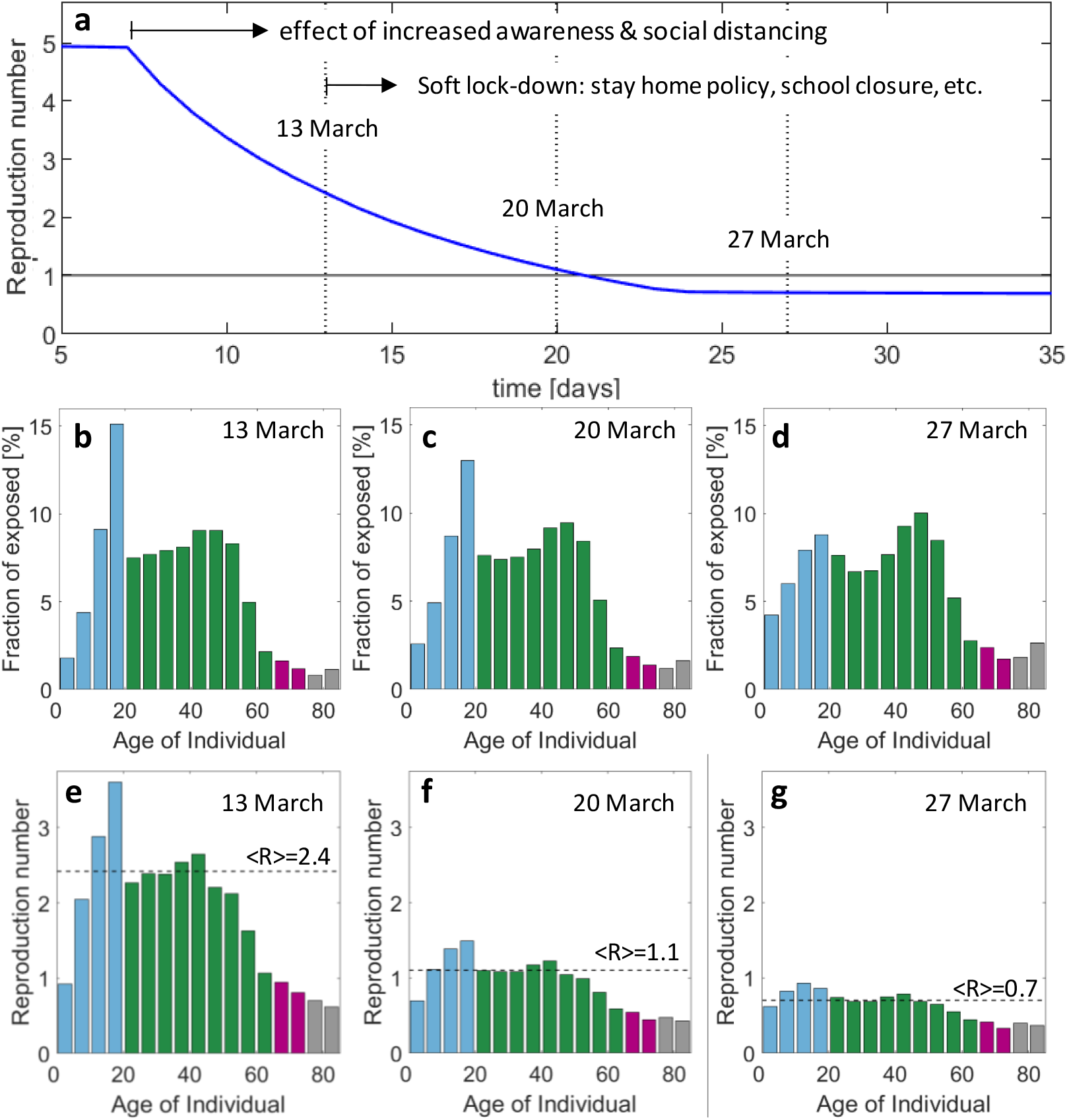
Reproduction number for first wave of epidemic. **(a)** Evolution of reproduction number (average weighted by the age distribution of the exposed) during transient phase induced by increased awareness and government measures. Age distribution of the compartment of exposed individuals on **(b)** March 13, **(c)** March 20 and (d) March 27. The corresponding age-stratified reproduction numbers for these dates are shown in **(e)-(g)**.

**Fig 4.**
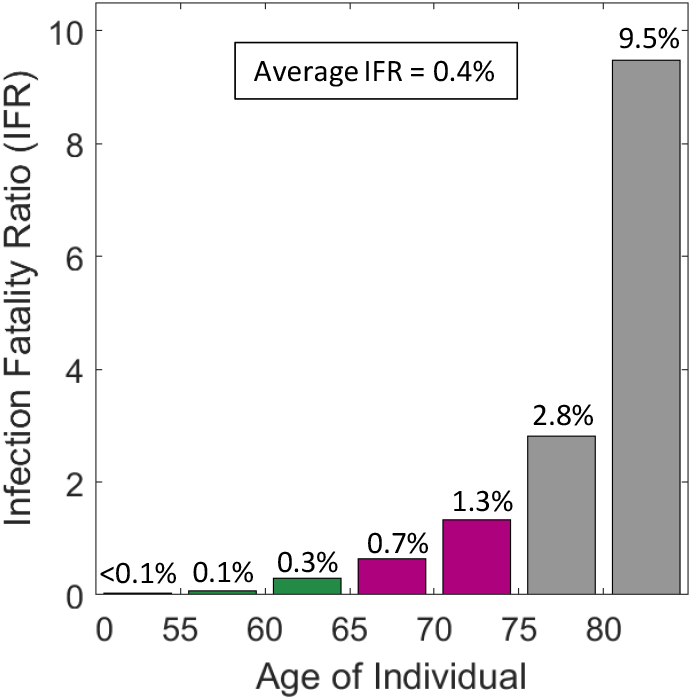
Estimated age-specific infection fatality ratio (IFR) The weighted average is computed based on the age distribution in the compartment of exposed individuals on April 20^th^.

To elucidate the importance of non-pharmaceutical intervention measures, we simulated the progression of the epidemic using the contact matrices and transmission probabilities that characterize the situation shortly before the lockdown (average reproduction number of 2.4). For this hypothetical scenario, herd immunity would have been reached within 2 to 3 months (Figs. 5a-c). By May 15, about 75% of the population would have been infected with an approximately uniform fraction of infected of about 85% in the 20 to 55 years age-groups (Fig. 5a). Close to 97% of the 15-20 year old would have been infected, while less than 50% would have been exposed to the coronavirus among those older than 65 years. The associated peak ICU demand for COVID-19 patients would have been close to 60 beds per 100’000 capita (Fig. 5b). The total of fatalities plateaus at about 22’000 with a loss of about 0.9% and 3% of the age-groups 75-79 and ≥80 years (Fig. 5c). At the opposite extreme, we simulated the scenario where the extraordinary state is maintained without any relaxation of measures. Less than 10% of the population would have been infected by the end of 2020 (Fig. 5d), while the peak in ICU need would have attained about 5 beds per 100’000 (Fig. 5e). The total of fatalities remains below 2,300 with less than 0.4% of those aged 80 years and older losing their life because of COVID-19 (Fig. 5f).

**Fig 5.**
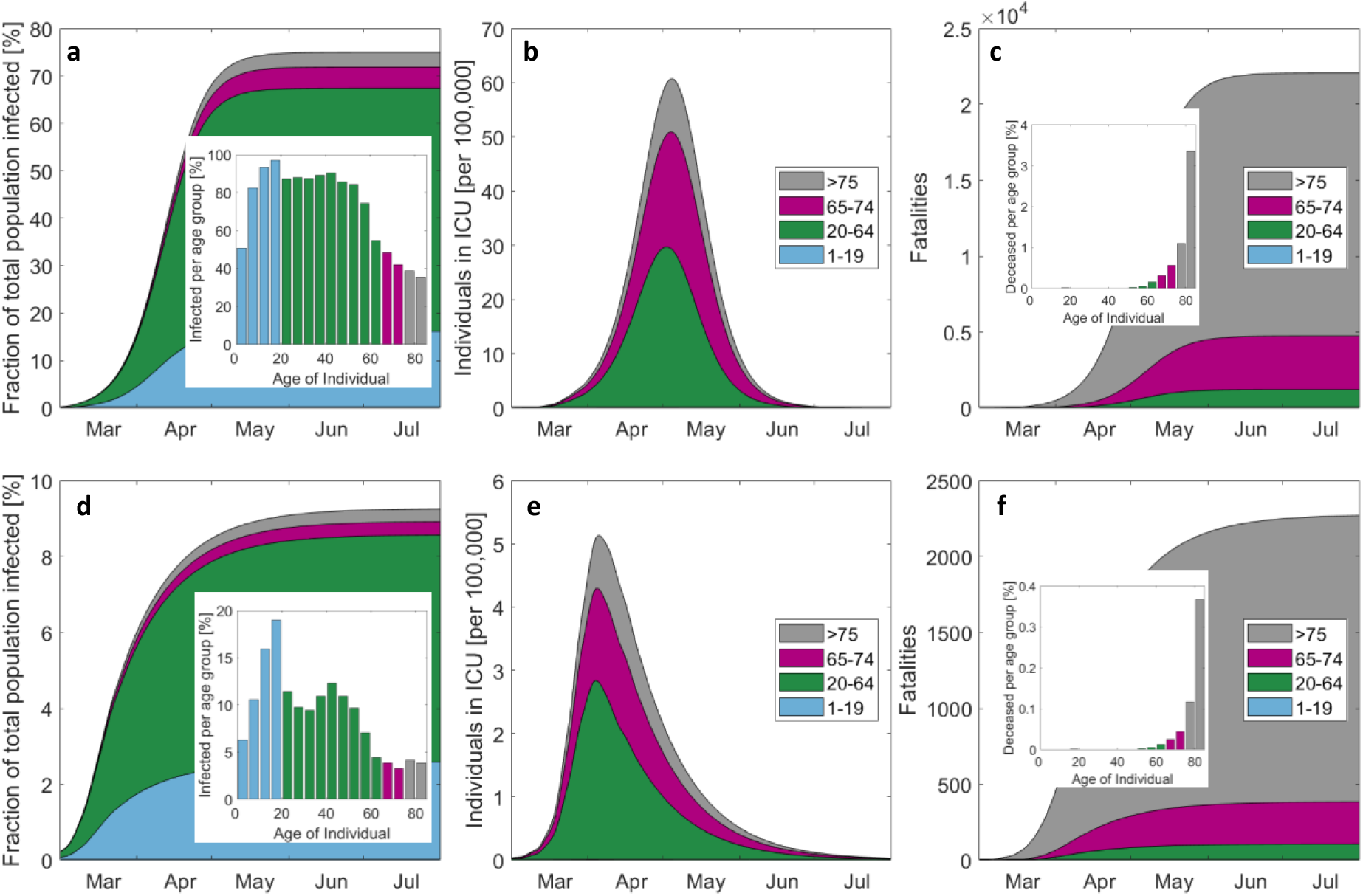
Evolution of the epidemic for the hypothetical cases of no measures taken (top) and perpetual extraordinary state (bottom) **(a)** and **(d)** show the cumulative cases and the final age-distribution by the end of the epidemic, the number of individuals in ICU is shown by **(b)** and **(e)**, while the cumulated number of fatalities is depicted in **(c)** and **(f)**. Note that the scales of the top and bottom figures differ by one order of magnitude.

After putting the country into an extraordinary state that guarantees an overall reproduction number well below 1, the goal is to ease the measures in a way that a “new-normal” state is attained, which allows for a well-functioning economy (i.e. businesses reopen with most people back to work). The main constraint is that a second wave of infections needs to be avoided or at least be recognizable at an early stage to take corrective measures. Selected elements of the plan proposed by the Swiss government are summarized in Fig. 6.

**Fig 6.**
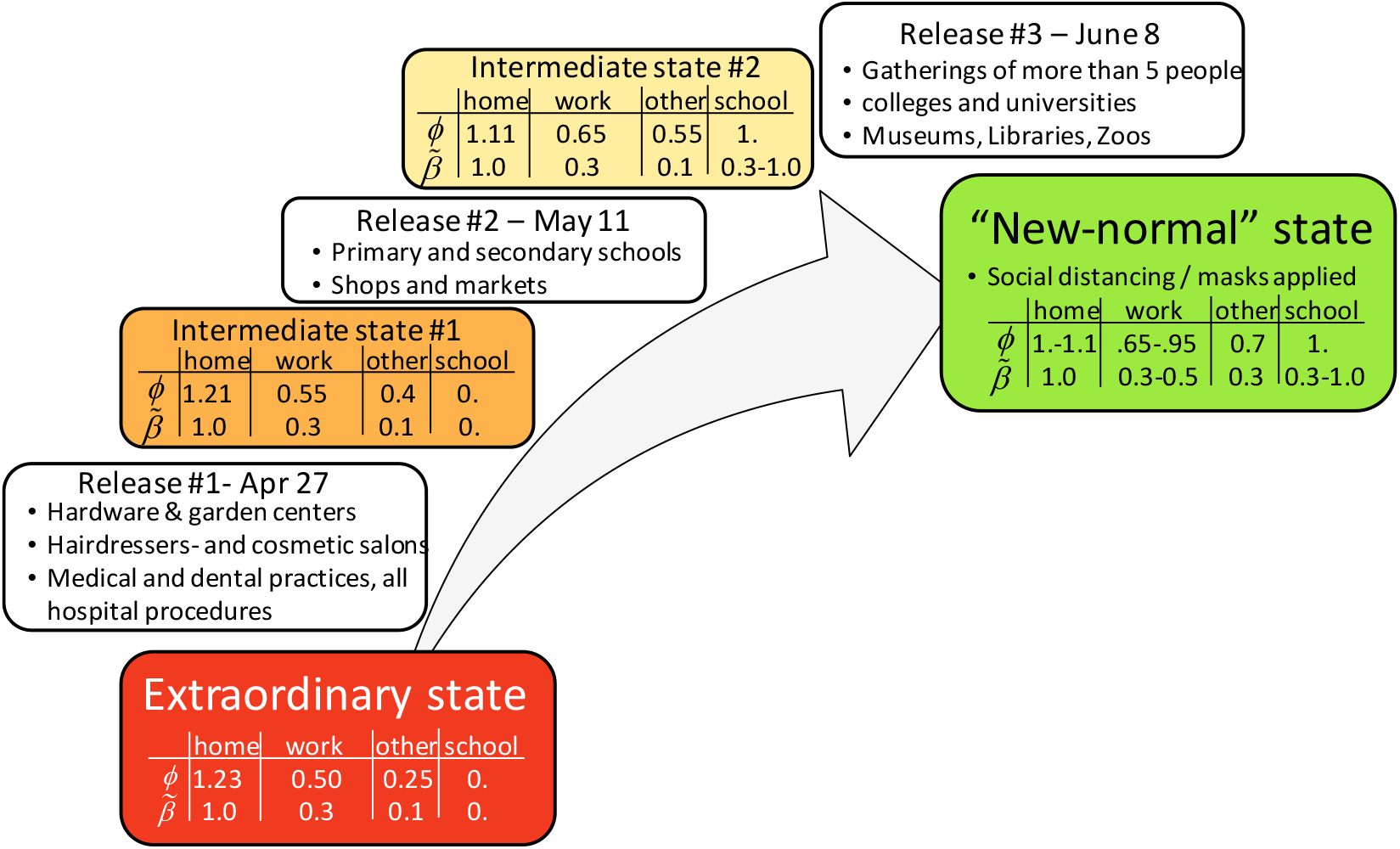
Roadmap for transitioning from an extraordinary state to a “new-normal” state. The progressive release will generate two intermediate states. The scaling factors *ϕ*_i_ for the location-dependent contact matrices and the associated knock-down factors *β*^*^ for the transmission probability prior to the soft lock-down are given in a table for each state.

In our model, we assume that two intermediate states are attained during the stepwise release of measures. In the absence of experimental data, assumptions regarding the modified contact patterns (as represented by the location-specific contact weighting factors *ϕ* and transmission probabilities *β*^*^ in Fig. 6) are made based on a combination of intuition and common practice in epidemic modeling. An important coupling is that the number of contacts at home decreases as the fractions of individuals at work and school increase. In a first scenario, we consider that (i) schools reopen without any special safety measures in place (by assuming the same probability of transmission before and after the lockdown, i.e. 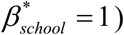 and that (ii) 65% of the workforce is physically present at work by June 8^th^. The main difference between the “new-normal” state and the state prior to the (soft) lockdown is that social distancing is enforced (or at least masks are worn) at work and other locations, that 30% of the workforce is in home office. The simulation results (Fig. 7a) suggest that the first release of measures will only have a minimal effect on the epidemic with a reproduction number staying well below 1 from April 27 to May 11.

**Fig 7.**
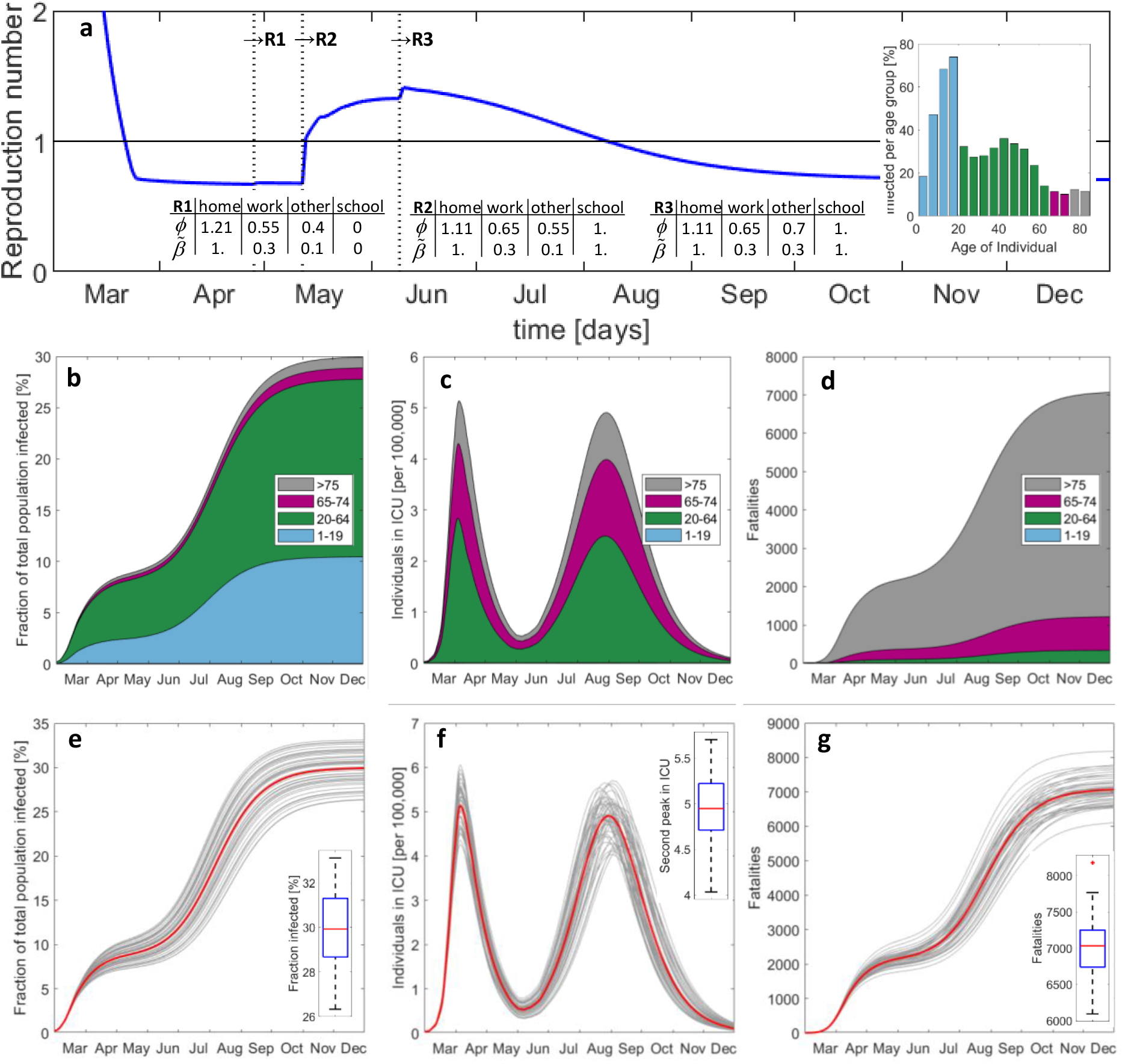
Predicted evolution of the epidemic after step-wise release of measures including school reopening without special caution. Histories of **(a)** average reproduction number, **(b)** total number of exposed, **(c)** individuals in ICU, and **(d)** accumulated fatalities. The vertical lines in (a) indicate the instants of release measures #1 to #3. The inserted bar plot shows the infected per age-group by December 2020. **(e)-(g)** Results from Monte-Carlo analysis. The 50 gray curves in each plot are obtained after a random perturbation of the main model parameters, while the red curve corresponds to the respective deterministic solution reported in (b)-(d). The boxplots show the median (red), the 25th and 75th percentiles (in blue), the extreme data (black) and outliers (red star symbol).

However, the second release (which involves the reopening of schools) is expected to cause a second peak of infections (Fig. 7b), generating approximately the same daily hospital and ICU load by mid-August as the first peak (Fig. 7c). The second peak is less steep than the first. As a result, there is a two-month time window from early June to the end of July where the second peak could be detected through the monitoring of the increase of the hospitalizations. In other words, there would be less pressure on the government to take additional corrective measures quickly than at the beginning of the epidemic in March. However, this reasoning only applies when focusing on preventing the collapse of the healthcare system. The increased width of the second peak also implies a significantly higher death toll. While the first wave of infections generated about 2,000 fatalities, the second wave could generate almost 5,000 additional deaths (Fig. 7d). It is thus extremely dangerous if the virus spreads with a reproduction number slightly higher than 1. The public perception may be positive as long as the healthcare system can handle the load of ICU patients, while many would silently die until reaching herd immunity. In the current scenario, herd immunity would be achieved by the end of 2020 after infecting about 30% of the total population. To gain insight into the robustness of the model predictions, a Monte-Carlo analysis is performed where we introduced six multipliers to perturb the probabilities of (i) developing symptoms, (ii) hospitalization, (iii) transfer to ICU, and (iv) death in self-isolation, (v) in MCU, and (vi) in ICU. Assuming a uniform distribution of each multiplier over an interval [0.9,1.1] (i.e. a standard deviation of 5.7%), 50 simulations were run with randomly drawn multipliers. The results shown in Figs. 7e-g reveal standard deviations (normalized by their means) of 8.6%, 6.0% and 5.6% for the second peak in ICU need, for the population infected and for the fatalities, respectively. Among the model parameters, the highest sensitivity of the number of deaths is observed for variations in the probability of hospitalization.

To achieve a more positive outcome of the release of measures, we redid the above simulations with reduced probabilities of transmission at school (Fig. 8). It turns out that the overall reproduction number after reopening schools will remain below 1 if the probability of transmission (per contact at school) is reduced by 50%. In that case, a second peak could be avoided. Moreover, and probably most importantly, the excess fatalities associated with school reopening would drop from 5,000 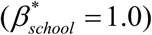 to less than 1,000 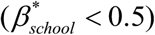. Any failure of maintaining the rate of transmission at school reasonably low would result in a substantial increase in fatalities within a few months to a level that is significantly higher than the annual total of fatalities associated with influenza (about 1,000 for Switzerland). Repeating the above simulations with the assumption that 95% of the workforce is physically present at work (which also increases the probability of transmission at work from 0.3 to 0.5) resulted in similar results (Supplementary Fig. 5). In the Supplementary Information, we also considered the immoral (and practically-infeasible) scenario of temporarily isolating individuals older than 70 years from the rest of the society while letting life resume for all other age-groups without any restrictions (with an initial reproduction number of about 2.4). For that scenario, the total death toll would be 4,100 before reaching herd immunity (67% of total population infected) within two months (Supplementary Fig. 6). However, the peak hospital and ICU demand of 29 beds (per 100’000) are likely to exceed the capacity which may cause additional fatalities for this scenario.

**Fig 8.**
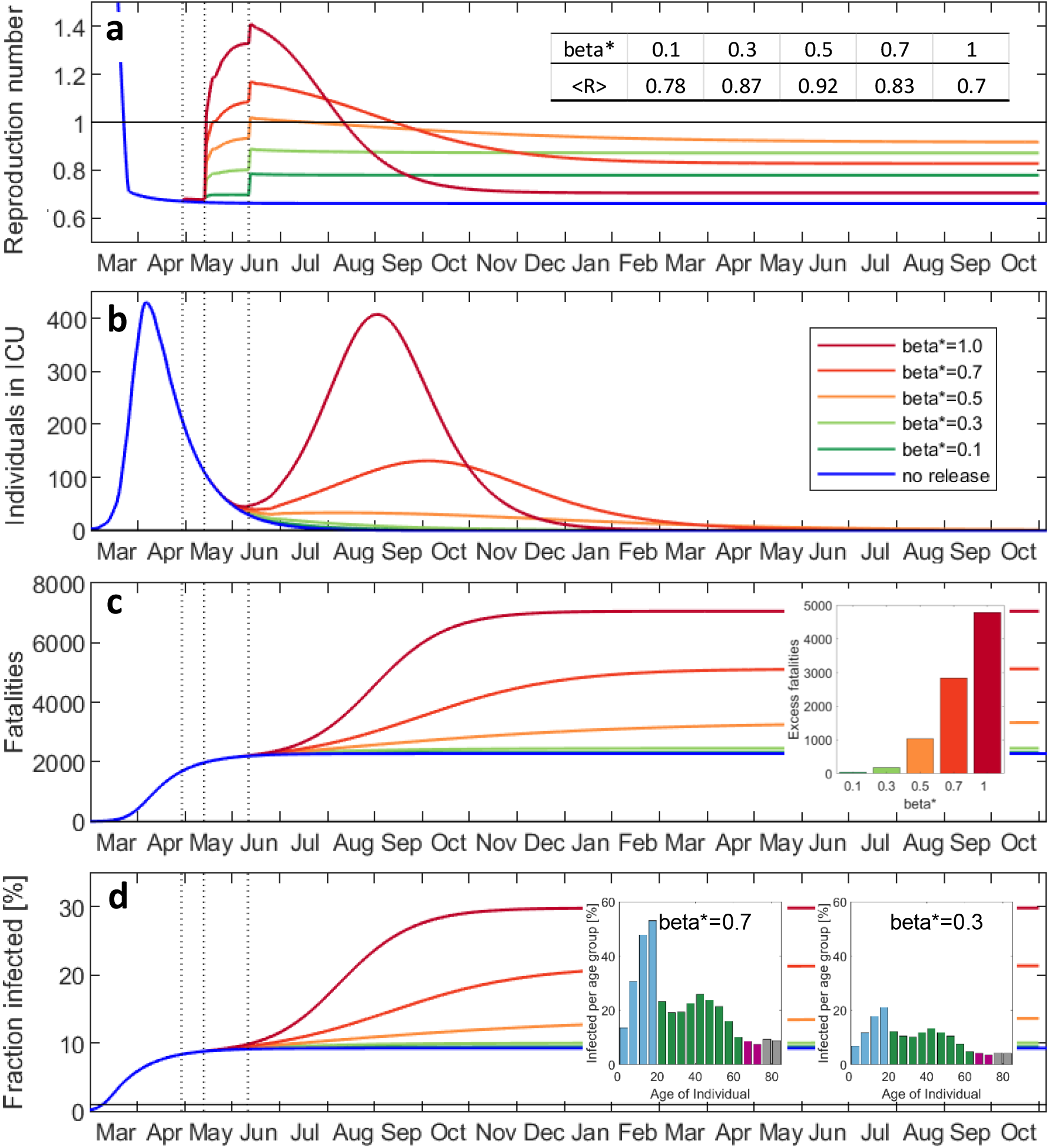
Effect of special caution at school on the evolution of the epidemic. Histories of **(a)** average reproduction number, **(b)** individuals in ICU, **(c)** accumulated fatalities, **(d)** fraction of the total population infected. The factor *β\** represents the reduction of the probability of transmission at school. The table insert in (a) denotes the average reproduction number in October 2021. The insert in (c) shows the death toll (fatalities in excess of result for no release) for reopening schools as function of *β*^*^. The inserts in (d) illustrate the age-distribution of the infected.

The accuracy of the model predictions hinges on the availability of high quality data and the understanding of the transmission dynamics of the SARS-CoV-2 virus. Both elements are expected to improve over time which requires the constant updating of the model. On the data side, the combined antibody and PCR-testing of a random group of about 10’000 individuals (Supplementary Information) is expected to provide valuable improvements of the assumptions made regarding the probabilities of hospitalization and the fraction of the asymptomatic group. The transport and transmission of COVID-19 through children is also unclear at this stage. When repeating the release scenario simulations with *β_school_* = 0.3 from age 10 upwards, while maintaining *β_school_* = 1 for those aged 10 years or younger, we obtain approximately the same response as for *β_school_* = 0.3 for all age-groups (Supplementary Fig. 7). This result suggests that the risk associated with reopening kindergardens and elementary school might be worth taking even if (i) future research demonstrates that kids are as infectious as other age-groups, and if (ii) safety measures (such as wearing masks) at school are not implemented for those aged <10 years. The analysis of the multi-variate effect of the probabilities of transmission at school, work and other locations (Supplementary Information) revealed that effective protection measures at all three locations are crucial for limiting the total of fatalities.

In summary, a new discrete modeling framework is proposed to capture the dynamics of highly age-sensitive epidemics and to evaluate the effect of social contact patterns on the load of hospitals and their intensive care units. The model architecture is specified to describe the COVID-19 epidemic before identifying all model parameters based on the data reported for Switzerland. It is demonstrated that the model provides an accurate description of the history and the age-distributions of the individuals hospitalized, in ICU and deceased. The obtained (low) infection fatality ratio of 0.4% for Switzerland is in agreement with the testing-based estimate for the German county Heinsberg^19^. The model-based analysis of the outbreak elucidates the highly non-uniform attack of COVID-19. It is estimated that the reproduction number for the mostly highly-infected age-group (15-20 years old) is up to five times higher than that of the least infected age-group (>80 years) which appears to be protected ‘naturally’ by the social contact patterns. Nonetheless, individuals aged 80 and higher make up for 70% of the fatalities as opposed to less than 0.1% for those below 50 years. Simulations of the three-phase lifting of the soft lockdown in Switzerland demonstrate the need to reduce the probability of transmission at school (even when assuming an effective protection at all other locations) to avoid a second wave of COVID-19 infections. Given that the reproduction number is much lower during the possible build-up of the second peak in hospitalizations (as compared to the first peak), there should be sufficient time for governments to take corrective measures in case they detect a significant increase in hospitalizations after relaxing the lockdown. Even though the second wave may not lead to a collapse of the healthcare system, it is still important to maintain the overall reproduction number always below unity to avoid the silent multiplication of the total number of fatalities. Most parameters and features of the detailed COIVID-19 model for Switzerland will also be highly relevant for forecasting the effect of lockdown relaxation measures in other countries. By constantly training the proposed epidemic model based on current data for several countries, it will also be possible to quantify the effect of government measures on the rate of contact and transmissions from real-life observations. This important insight will not only be relevant for managing the current COVID-19 crisis, but also improve the reliability of predictions in the event of future pandemics.

## Methods

### Data

We consider data sets collected by the private platform www.corona-virus.ch after sporadically cross-checking the data sets with those provided by official sources (Swiss Federal Office of Health (BAG) and the canton Vaud). Information on age-distributions is also obtained from the official sources. The age-distribution in Switzerland is obtained from data for 2016 reported on www.populationpyramid.net (Supplementary Fig. 2).

### Epidemic model

The population is stratified into n=17 age-groups: #1 (0-4), #2 (5-9), …, #16 (75-79) and #17 (>80). In addition, the population is partitioned into nine compartments: susceptible (S), asymptomatic infectious (A), symptomatic infectious (B), symptomatic in self-isolation (C), hospitalized in middle care unit (H), hospitalized in intensive care unit (Q). Furthermore, each compartment is divided into sub-compartments which classify the individuals according to the number of days spent in a given compartment. The state of each sub-compartment is represented by a vector whose components correspond to the number of individuals for a specific age-group. For example, the third component *C*_3_^(4)^ of the vector **C^(4)^** provides the number of symptomatic individuals of age-group #3 (10-14 years) that are spending their fourth day in self-isolation. The time resolution of the model is fixed to Δ*t* = 1 *day*. The main modeling assumptions are:

- Symptomatic individuals are 50% more infectious than asymptomatic individuals^1^. Even though the viral loads in symptomatic and asymptomatic patients appear to be similar^7^, the higher probability of infecting through coughing supports the assumption of a higher infectiousness for the symptomatic patients.
- The new infections (i.e. individuals in sub-compartment **E**^(1)^) are then given by

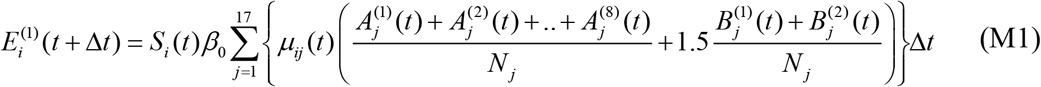

with the probability of transmission *β*_0_ (per contact) for March 13, 2020 (day of soft lock-down) and the effective contact matrix (contacts per day)

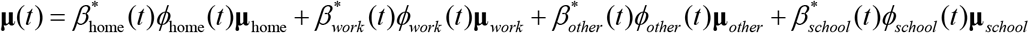

where the dimensionless multipliers *β\** and *ϕ* account for the location-dependent changes in the transmission probability and contact frequency due to non-pharmaceutical intervention measures. The contact matrices describing the contact patterns at home (**μ**_home_), at work (**μ***_work_*\*), at school (**μ***_school_*,) and at other locations (**μ***_other_*) are shown in Supplementary Fig. 2. They have been obtained for Switzerland from Perm et al.^20^, extended for the age-group 80+ and readjusted to satisfy reciprocity for the assumed age-distribution.
- The incubation time is fixed to 5 days which is close to the mean incubation period of 5.2 days (with a 95% confidence interval of 4.1-7.0) observed in Wuhan^21^.
- From the reports of the testing of all 3,711 passengers of the cruise liner Diamond Princess^13^ and of 85% of the population of the Italian municipality Vo’ (2,812 subjects)^14^, it is assumed that *λ_SYM_* = 60% of the infected develop symptoms.
- Based on the results from live virus isolation from the sputum of hospitalized cases in Germany^22^ and indications of similar viral loads in symptomatic and asymptomatic subjects in Italy^14^, it is assumed that asymptomatic individuals remain infectious for 8 days. Symptomatic individuals are expected to self-isolate two days after the onset of symptoms.
- Symptomatic individuals are assumed to transfer to hospital on their 5^th^ day of self-isolation. This assumption is made to match reports from international media that it takes approximately two weeks from first exposure to hospitalization.
- The middle care unit (MCU) features seven sub-compartments to match the average duration of hospitalization (outside ICU) of 7 days reported by the Swiss canton Vaud.
- The same canton also reported an average duration of a stay in ICU of 6 days. Due to the temporal distribution of the high number of deaths in ICU, the intensive care unit features eight sub-compartments in attempt to match the average duration in ICU of 6 days after accounting for deaths.
- The age-dependent transfer functions *λ_HOSP_* (from self-isolation to hospital) and *λ_ICU_* (from MCU to ICU) are inferred from the modeling of the outbreak in Switzerland (see main text). The same applies to the age-aggregated probabilities of death in self-isolation, MCU and ICU. These functions are depicted in Fig. 1. The corresponding numerical values are provided in Supplementary Tables 1 and 2.

Different from differential equation-based SIR-type models, there is no need to employ a numerical solver. After evaluating the algebraic equation (M1), the state of the model is updated by shifting the individuals sequentially from a first sub-compartment to a subsequent sub-compartment (Fig. 1) while applying the above transfer functions.

### Reproduction number

The reproduction number **R** is a vector whose components *R_j_* estimate the number of individuals (among all age-groups) that an infectious individual of age-group *j* would infect during its period of communicability (8 days for asymptomatic, and 2 days for symptomatic individuals). Assuming that the effective contact matrix and number of susceptible individuals remains constant during the period of communicability, it can be given by the approximation

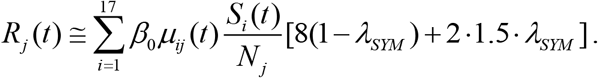

The overall reproduction number < *R >* is computed as weighted average using the current age-distribution of the exposed as weighting function.

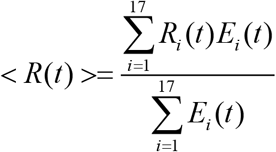

with 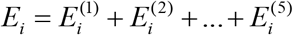.

### Infection Fatality Ratio (IFR)

Denoting the daily probabilities of death in self-isolation, MCU and ICU as 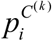, 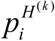 and 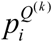, the probability of survival *(ISR)_i_* of an infected individual of age-group *i* reads

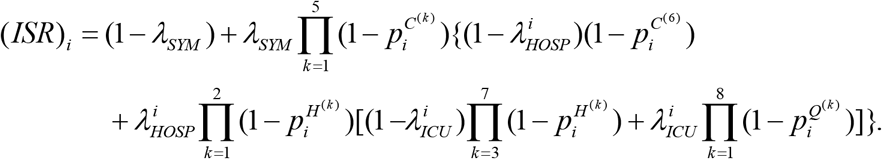

Hence, we have the corresponding IFR for age-group i,

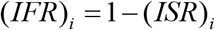

As for the reproduction number, we make use of the age-distribution for the exposed to calculate the average IFR.

## Data Availability

N/A

## Code availability

The codes are available upon request to the corresponding author.

## Author contributions

Both authors contributed equally to the work.

## References

1. Guan, W. et al. (2020), Clinical Characteristics of Coronavirus Disease 2019 in China, New England Journal of Medicine, 382:1708–20. DOI: https://doi.org/10.1056/NEJMoa2002032

2. Hethcote, HW (2000). The mathematics of infectious diseases, SIAM REVIEW 42 (4), DOI: https://doi.org/10.1137/S0036144500371907

3. Ferguson, N.M. et al. (2020). Impact of non-pharmaceutical interventions (NPIs) to reduce COVID-19 mortality and healthcare demand (preprint), DOI: https://doi.org/10.25561/77482.

4. Lin, Q., et al. (2020). A conceptual model for the coronavirus disease 2019 (COVID-19) outbreak in Wuhan, China with individual reaction and governmental action. International Journal of Infectious Diseases, 93, 211–216. https://doi.org/10.1016/Miid.2020.02.058

5. Prem, K., et al. (2020). The effect of control strategies to reduce social mixing on outcomes of the COVID-19 epidemic in Wuhan, China: a modelling study. The Lancet 261–270. https://doi.org/10.1016/S2468-2667(20)30073-6

6. Massonnaud, C., Roux, J., Crepey, P. (2020). COVID-19: Forecasting short term hospital needs in France (medRxiv preprint), DOI: https://doi.org/10.1101/2020.03.16.20036939

7. Wu, J.T. et al. (2020), Estimating clinical severity of COVID-19 from the transmission dynamics in Wuhan, China. Nat Med 26, 506–510. https://doi.org/10.1038/s41591-020-0822-7

8. Yeo, Yao-Yu, Yeo, Yao-Rui, Yeo, Wan-Jin (2020). A Computational Model for Estimating the Progression of COVID-19 Cases in the US West and East Coasts (medRxiv preprint), DOI: https://doi.org/10.1101/2020.03.24.20043026

9. Wu, J. T., Leung, K., & Leung, G. M. (2020). Nowcasting and forecasting the potential domestic and international spread of the 2019-nCoV outbreak originating in Wuhan, China: a modelling study. The Lancet, 395(10225), 689–697. https://doi.org/10.1016/S0140-6736(20)30260-9

10. Zhao, S., & Chen, H. (2020). Modeling the epidemic dynamics and control of COVID-19 outbreak in China. Quantitative Biology, https://doi.org/10.1007/s40484-020-0199-0

11. Giordano, G., Blanchini, F., Bruno, R., Colaneri, P., Filippo, A. Di Matteo, A. Di, & Colaneri, M. (2020). Modelling the COVID-19 epidemic and implementation of population-wide interventions in Italy, Nature medicine Letters, DOI: https://doi.org/10.1038/s41591-020-0883-7

12. Riou, J., Hauser, A., Counotte, M.J., Althaus, C.L. (2020). Adjusted age-specific case fatality ratio during the COVID-19 epidemic in Hubei, China, January and February 2020 (preprint), DOI: https://doi.org/10.1101/2020.03.04.20031104

13. Japanese National Institute of Infectious Diseases. Field Briefing: Diamond Princess COVID-19 Cases (2020) https://www.niid.go.jp/niid/en/2019-ncov-e/9407-covid-dp-fe-01.html

14. Lavezzo, E. et al. (2020). Suppression of COVID-19 outbreak in the municipality of Vo, Italy (medRxiv preprint), DOI: https://doi.org/10.1101/2020.04.17.20053157

15. Gudbjartsson, D.F. et al. (2020). Spread of SARS-CoV-2 in the Icelandic Population. The New England Journal of Medicine, https://doi.org/10.1056/NEJMoa2006100

16. Intensive Care National Audit & Research Centre (ICNARC), ICNARC report on COVID-19 in critical care, 17 April 2020, www.icnarc.org

17. Verity, R. et al. (2020). Estimates of the severity of coronavirus disease 2019: a model-based analysis. Lancet, 1–9, DOI: https://doi.org/10.1016/S1473-3099(20)30243-7

18. Scire, J., et al. (2020). Reproductive number of the COVID-19 epidemic in Switzerland with a focus on the Cantons of Basel-Stadt and Basel-Landschaft, Swiss Medical Weekly, DOI: https://doi.org/10.4414/smw.2020.20271

19. Streeck, H., et al. (2020). Infection fatality rate of SARS-CoV-2 infection in a German community with a super-spreading event. Available at https://www.ukbonn.de/42256BC8002AF3E7/direct/home

20. Prem, K., Cook, A. R., & Jit, M. (2017). Projecting social contact matrices in 152 countries using contact surveys and demographic data. 1–21. https://doi.org/10.1371/journal.pcbi.1005697

21. Li, Q. et al. (2020). Early Transmission Dynamics in Wuhan, China, of Novel Coronavisus-Infected Pneumonia, New England Journal of Medicine 382 (13), 1199.

22. Wölfel, R. et al. (2020). Virological assessment of hospitalized cases of coronavirus disease 2019 (medRxiv preprint), DOI: https://doi.org/10.1101/2020.03.05.20030502

